# National Early Warning Scores and COVID-19 deaths in care homes: a longitudinal ecological study

**DOI:** 10.1101/2020.06.15.20131516

**Authors:** Daniel Stow, Robert O Barker, Fiona E Matthews, Barbara Hanratty

## Abstract

**Objectives:** To investigate whether patterns of National Early Warning Scores (NEWS/NEWS2) in care homes during the COVID pandemic correspond with area-level COVID-19 death registrations from care homes.

**Study design:** Longitudinal ecological study.

**Setting:** 460 Care home units using the same software package to collect data on residents, from 46 local authority areas in England.

**Participants:** 6,464 care home residents with at least one NEWS recording.

**Exposure measure:** 29,656 anonymised person-level NEWS from 29/12/2019 to 20/05/2020 with component physiological measures: systolic blood pressure, respiratory rate, pulse rate, temperature, and oxygen saturation. Baseline values for each measure calculated using 80^th^ and 20^th^ centile scores before March 2020.

**Outcome measure:** Time series comparison with Office for National Statistics (ONS) weekly reported registered deaths of care home residents where COVID-19 was the underlying cause of death, and all other deaths (excluding COVID-19) up to 10/05/2020.

**Results:** Deaths due to COVID-19 were registered from 23/03/2020 in the study geographical areas. Between 23/03/2020 and 10/05/2020, there were 5,753 deaths (1,532 involving COVID-19 and 4,221 other causes). The proportion of above-baseline NEWS increased from 16/03/2020 and closely followed the rise and fall in COVID-19 deaths over the study period. The proportion of above-baseline oxygen saturation, respiratory rate and temperature measurements also increased approximately two weeks before peaks in care home deaths in corresponding geographical areas.

**Conclusions:** NEWS may make a useful contribution to disease surveillance in care homes during the COVID-19 pandemic. Oxygen saturation, respiratory rate and temperature could be prioritised as they appear to signal rise in mortality almost as well as total NEWS. This study reinforces the need to collate data from care homes, to monitor and protect residents’ health. Further work using individual level outcome data is needed to evaluate the role of NEWS in the early detection of resident illness.

## Introduction

Care homes have experienced high rates of COVID-19 infection and death. In England, over half of excess mortality in the first months of 2020 is estimated to have been in this setting.^1^ Surveillance of COVID-19 in care homes has been difficult, because of a paucity of testing, and the lack of experience with how this disease presents in older people.^2^

An increasing number of UK care homes are collecting data on the National Early Warning Score (NEWS). This is a ‘track and trigger’ system used in hospitals in the United Kingdom to identify patients at risk of acute deterioration. NEWS requires the measurement of six parameters: temperature, pulse, systolic blood pressure, respiratory rate and oxygen saturation, along with conscious level. The score generated should trigger a pre-specified response, ranging from repeating the score within a specific timeframe, to seeking urgent medical attention.^3^ The British Geriatric Society produced guidelines for managing COVID- 19 in care homes for older people. To support triage, they recommend training staff to measure a resident’s vital signs (temperature, blood pressure, heart rate, pulse oximetry and respiratory rate) when COVID-19 infection is suspected.^4^ The risks and benefits of this approach are unknown. In community settings, elevated NEWS scores have been associated with prompt review by a health professional and poor health outcomes.^5^ Evidence to support the use of NEWS in care homes is limited, and whether it can support care home staff to identify or predict deterioration in the health of residents is unclear.^67,8^ NEWS observations, particularly measurement of blood pressure, require close contact between residents and care home workers, which may transmit COVID-19 infection.

This study will address the dearth of published evidence on the use of NEWS in any setting during the COVID-19 pandemic.^9^ The aim is to describe change in NEWS and its components over time and align these data with COVID-19 and all-cause mortality in care homes in England. We address the question of whether NEWS can contribute to surveillance during the pandemic, and whether an abbreviated NEWS (excluding one or more of the component measures) would suffice.

## Methods

### Study design and setting

We conducted an ecological study using individual level data for the exposure of interest (NEWS) and area level aggregate data for the outcome of interest (all cause and COVID-19 mortality). Participants were all residents of care homes utilising the same commercial software with cloud storage of data, and they all had at least one NEWS recording.^10^ We did not apply any further population inclusion/exclusion criteria.

Anonymised person-level NEWS (and the slightly modified NEWS2) information were obtained from 01/12/2019-20/05/2020 including the individual component measures of NEWS: blood pressure, respiratory rate, pulse rate, temperature, and oxygen saturation. Demographic information on the care home resident (age and sex) were obtained, along with a geographical identifier to examine regional variation. Geographical death data were obtained from Office for National Statistics (ONS) weekly reported registered deaths in care homes due to COVID-19, and all cause deaths (excluding COVID-19) available from week 1 (beginning 29/12/2019), to week 19 (ending 10/05/2020).^11^ ONS reporting areas and care home geographical labels were mapped as closely as possible.

### Analysis

We established baseline levels for NEWS and its component observations, in our population using centile cut points in a random subset of 70% of data collected between 12/2019 and 01/03/2020 (prior to the likely outbreak of COVID-19 in care homes in England). These cut points were then validated in the remaining 30% of observations made before March 2020. NEWS scores above the 80^th^ centile score were defined as above-baseline. For individual physiological observations, upper and lower thresholds were established using the 80^th^ and 20^th^ centile measurements in the same random subset (we calculated the lowest 20^th^ centile only for oxygen saturation). These cut points represent markers of increase in each parameter at a population level and are not measures of individual clinical concern. We removed biologically implausible values from the dataset before creating the centile cut points (Supplemental Table 1). We used quantile regression to measure the impact of age and sex on centile scores but did not find evidence to support age/sex specific 20^th^ and 80^th^ centile cut points in this population (details available from the authors on request).

We calculated the proportion of above-baseline NEWS measurements and the component observations, on a daily and weekly basis across all geographical areas provided. We plotted the proportion of weekly above-baseline measures in participating care homes as a time series against the weekly number of care home deaths due to COVID-19 and all-cause mortality (excluding COVID-19) occurring in the geographical areas present in our data. We calculated the cross-correlation between above-baseline NEWS scores and daily deaths (all causes) for time lags between zero and seven weeks. The individual physiological observations that anticipated COVID-19 mortality trends were combined to see if these could be useful for collection instead of the full NEWS panel.

All data management and analyses were conducted using R version 3.6.3^12^ we used the ccf function in the stats R package to calculate cross-correlations.

## Results

### Care home population

Care home data were available from 6,464 individuals, 2,007 men (mean age 80.1 years, SD=12.6) and 3,373 women (mean age 83.0 years, SD=12.9). Information on gender was missing from 1,086 (16.8%) people, and age information was missing for 116 (1.8%) people.

### Geographical variation in reporting

29,656 NEWS recordings were made across 46 Local Authority (LA) areas, from 480 unique care home IDs (identifiers for the device used to record the measurement, representing a care home, or a distinct unit within a care home). Most recordings were made in two LAs in the north east of England (n=11,029 and n=10,347), and in one London borough (n=3,411).

### Deaths in care homes

There were 10,407 registered deaths in care homes in the 46 LA and CCG areas between 29/12/2019 and 10/05/2020. The first death from COVID-19 was registered in week commencing 23/03/2020. From 23/03/2020 to 10/05/2020, there were 5,753 deaths of care home residents - 1,532 with an underlying cause of COVID-19 and 4,221 due to causes excluding COVID-19.

### Baseline NEWS centile scores

Table 1 contains information on NEWS taken from 9,586 (70% subset of 13,694) recordings made before 01/03/2020. Table 1 also contains thresholds for NEWS 20^th^ centile scores, and 80^th^ and 20^th^ centile scores for NEWS components calculated in this subset, and the proportion of observations exceeding these values in the 30% validation dataset.

**Table 1:** NEWS values before March 2020 in the development dataset, cut points for baseline measurements and proportion of above baseline measurements in the validation dataset

| Measurement | Development dataset |  |  |  |  | Validation dataset |  |  |
| --- | --- | --- | --- | --- | --- | --- | --- | --- |
|  | median | min | max | IQR* | 20th centile | 80th centile | % below 20th centile | % above 80th centile |
| NEWS | 2.0 | 0.0 | 15.0 | 3.0 |  | >4 |  | 14.7% |
| Temperature (C) | 36.5 | 32.0 | 40.0 | 0.6 | <36.2 | >=36.9 | 18.4% | 25.8% |
| Pulse rate (beats/min) | 76.0 | 22.0 | 212.0 | 19.0 | <66 | >=89 | 19.6% | 20.0% |
| Systolic BP (mmHg) | 123.0 | 50.0 | 235.0 | 26.0 | <109 | >=143 | 20.5% | 19.4% |
| Respiratory rate (breaths/min) | 19.0 | 6.0 | 60.0 | 3.0 | <17 | >=22 | 19.4% | 18.3% |
| Oxygen saturation (%) | 95.0 | 47.0 | 100.0 | 4.0 | <=92 |  | 17.6% |  |

\*IQR - Interquartile range

### NEWS measurements and care home deaths

The proportion of above baseline NEWS observations was stable from week 1 (30/12/2019 – 05/01/ 2020) until week 12 (16/03/2020-22/03/2020) and week 13 (23/03/2020- 29/03/2020) when there was a marked increase. This increase happened in the weeks before the majority of COVID-19 and non-COVID-19 deaths began to occur (from week 15 (06/04 – 12/04). The proportion of above baseline NEWS scores peaked in week 15, before beginning to decline again from week 16 (13/04-19/04) onwards (figure 1). The highest correlation was observed for a two-week lag (r=0.82, p≤ 0.05, supplemental figure 1)

**Figure 1:**
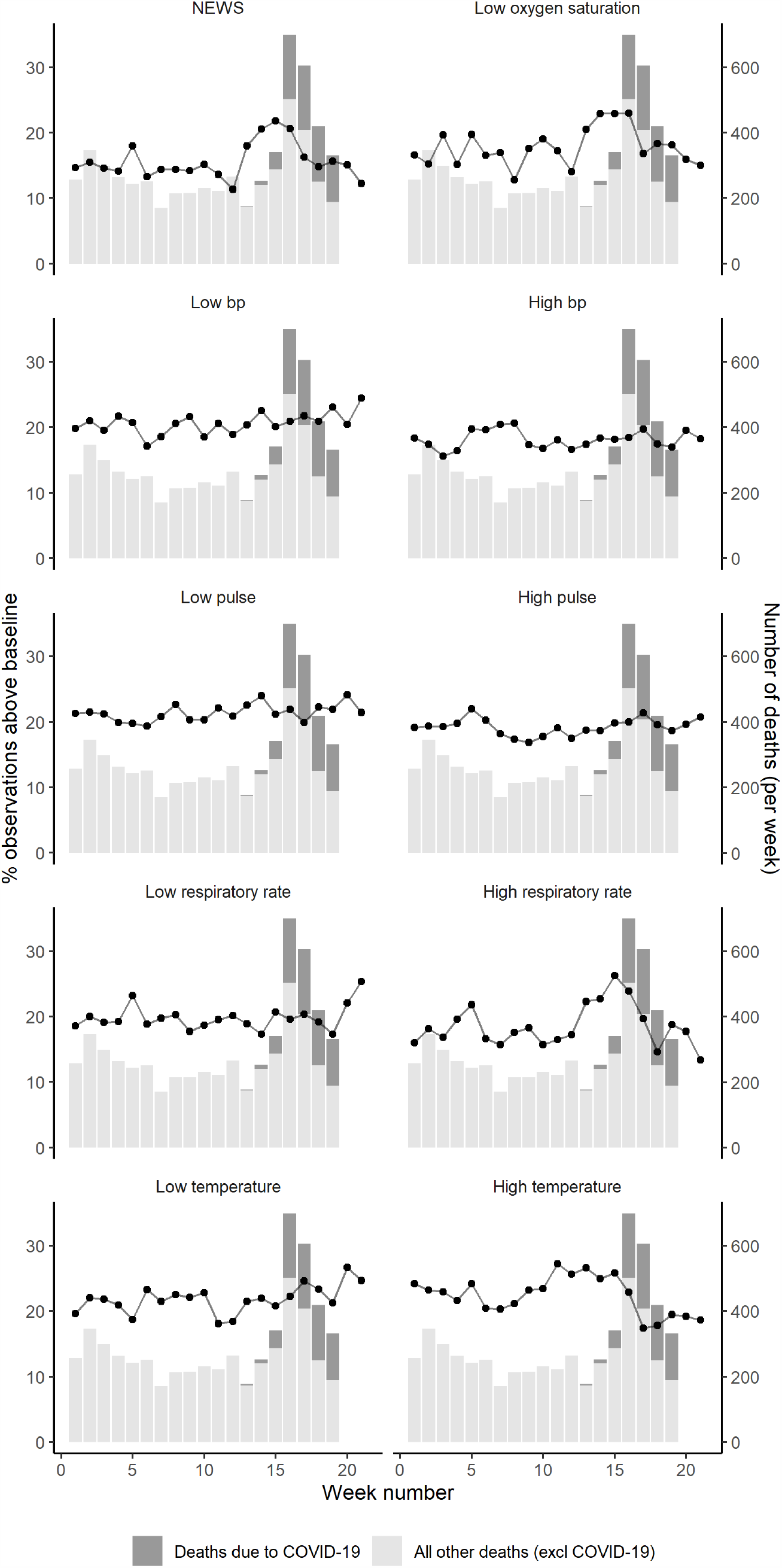
The proportion of above baseline NEWS and component measurements from December 2019 to May 2020 (lines), compared to care home deaths in corresponding geographical areas in England (bars).

### Individual NEWS component measures and care home deaths

The proportion of above baseline measures of high respiratory rate (r=0.73, p≤ 0.05 for a two-week lag) and low oxygen saturation (r=0.80, p≤ 0.05 for a two-week lag) appear to follow the pattern of COVID-19 and non-COVID-19 deaths more closely than other component measures (supplemental figure 1). The proportion of above baseline measures of temperature appeared to be decreasing between week 0 and week 10, before rising slightly to plateau until week 15 before declining again.

### Combination of NEWS component measures

Figure 2 shows paired combinations of above baseline respiratory rate and temperature and below baseline oxygen saturation. All increase just before peaks in COVID-19 and non COVID-19 deaths (Figure 2).

**Figure 2:**
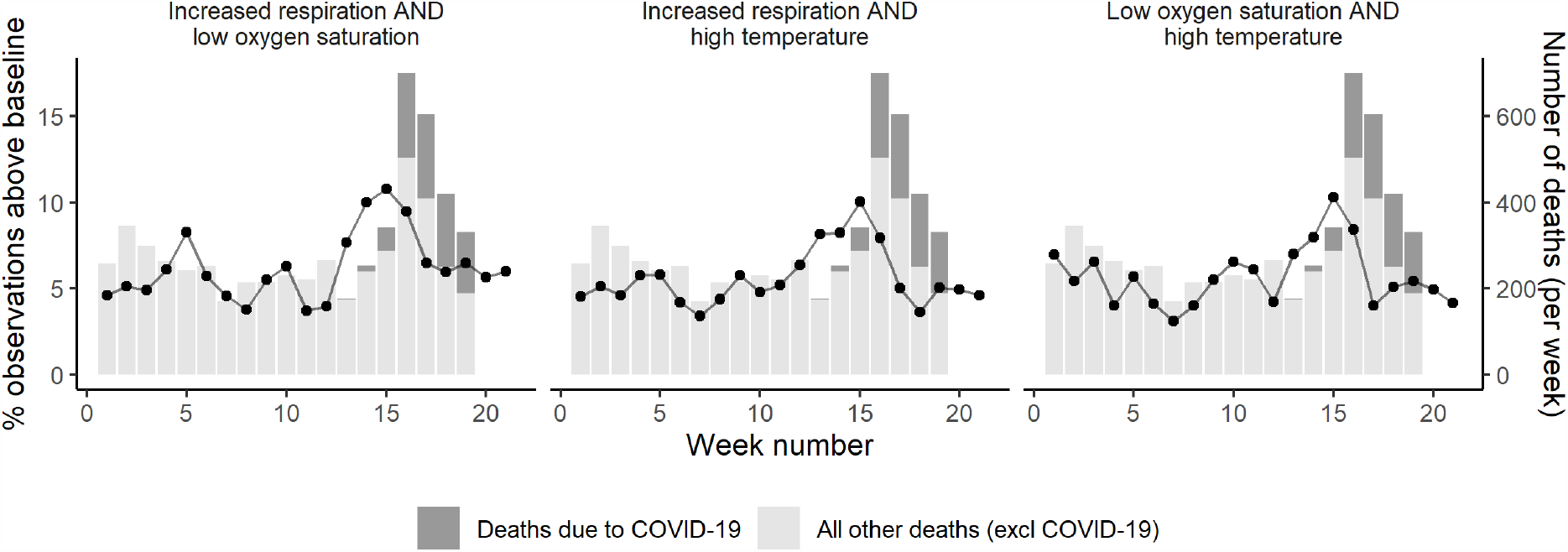
The proportion of above baseline NEWS component combinations December 2019 to May 2020, compared to care home deaths in corresponding geographical areas in England.

## Discussion

This study suggests that NEWS could make a useful contribution to disease surveillance in care homes during the COVID-19 pandemic. A rise in the proportion of above-baseline NEWS was observed from the middle of March 2020, when the incidence of COVID-19 was believed to be rising in the UK. The proportion of above-baseline measurements of oxygen saturation, respiratory rate and temperature also increased approximately two weeks before peaks in care home deaths in corresponding geographical areas. Oxygen saturation and respiratory rate appear to signal rise in mortality almost as well as total NEWS and may be safer and more practical to measure during a pandemic.

In this study, we observed a two-week time lag between peaks in NEWS measures and deaths. This is similar to the observed time between symptom onset and COVID-19 death in other settings.^13, 14^ Evidence for the role of NEWS in acute illness in community settings is growing,^15 16 17 18 19^ but to date, only one descriptive study has provided empirical data on NEWS in care homes.^6^ A recent systematic review on the use of NEWS in assessing unwell COVID-19 patients in primary care suggested that enthusiasm for its use may be premature.^9^ Our study does not provide support for the contribution of NEWS measurement to the care of individual residents, but it does suggest that it may have a role in health surveillance at a population level. Measurement and monitoring of some NEWS components may be useful in detecting future waves of COVID-19 infection in care homes. Emerging evidence suggests that up to half of care home residents do not have symptoms at the time they test positive for COVID-19.^20^ Whether NEWS measurement would detect approaching illness in any of these residents is unknown, and it cannot substitute for a comprehensive programme of testing in care homes.

### Strengths and limitations

To our knowledge, this is the first study to examine longitudinal variation in NEWS in care homes. We have described trends over time using a specific software system to collect data on NEWS in care homes. This means that the distribution of care homes within and between areas is not systematic, as it reflects the market share of the software company and local support for digital data collection in care homes. Most recordings were drawn from the north east of England, and a London borough, but we have no information on the proportion of care home residents in each area that are represented in our dataset. All the data were anonymised, and without individual outcome data, we examined patterns in and simple correlations between NEWS and area-level weekly registered death information. This study design was a pragmatic approach that made best use of available data, but it is not a causal study, nor a study of diagnostic accuracy, and it is liable to the ecological fallacy.

## Conclusions

NEWS may be useful to monitor health in care homes during the pandemic, but use of a shortened NEWS could be recommended. Oxygen saturation, respiratory rate and temperature provide a similar signal to the complete NEWS. The omission of some components of NEWS, such as blood pressure, minimises contact time between residents and care home staff, potentially reducing infection risk. Data on mortality and diagnoses, linked to NEWS, is required to evaluate the role of NEWS in assessing individual residents with suspected COVID-19 infection. Collection and aggregation of data from care homes would facilitate disease surveillance; we argue that introduction of a care home minimum dataset should be a priority for the UK.

## Data Availability

Data sharing: Information on the deaths of care home residents is feely available from the source cited.

## Ethical approval

This study was approved by a University Research Ethics Committee (Ref. 3297/2020) and was conducted under the Secretary of State’s directions under the Control of Patient Information Regulations, following advice from the Health Research Authority.

## Supplemental information

**Supplemental Table 1:**
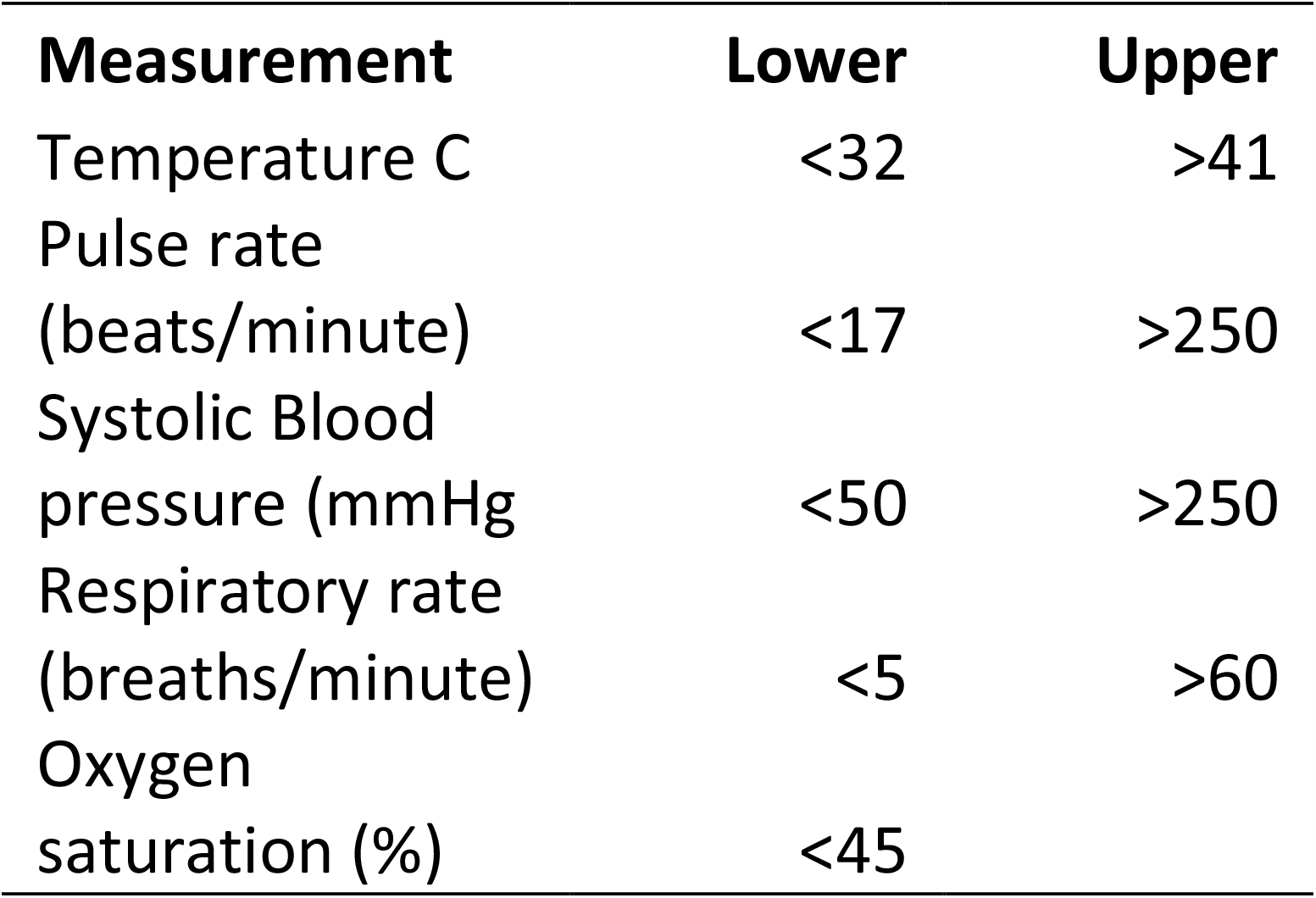
Filter values for biologically implausible NEWS component measurements

| Measurement | Lower | Upper |
| --- | --- | --- |
| Temperature C | <32 | >41 |
| Pulse rate<br>(beats/minute) | <17 | >250 |
| Systolic Blood<br>pressure (mmHg) | <50 | >250 |
| Respiratory rate<br>(breaths/minute) | <5 | >60 |
| Oxygen<br>saturation (%) | <45 |  |

**Supplemental Figure 1:**
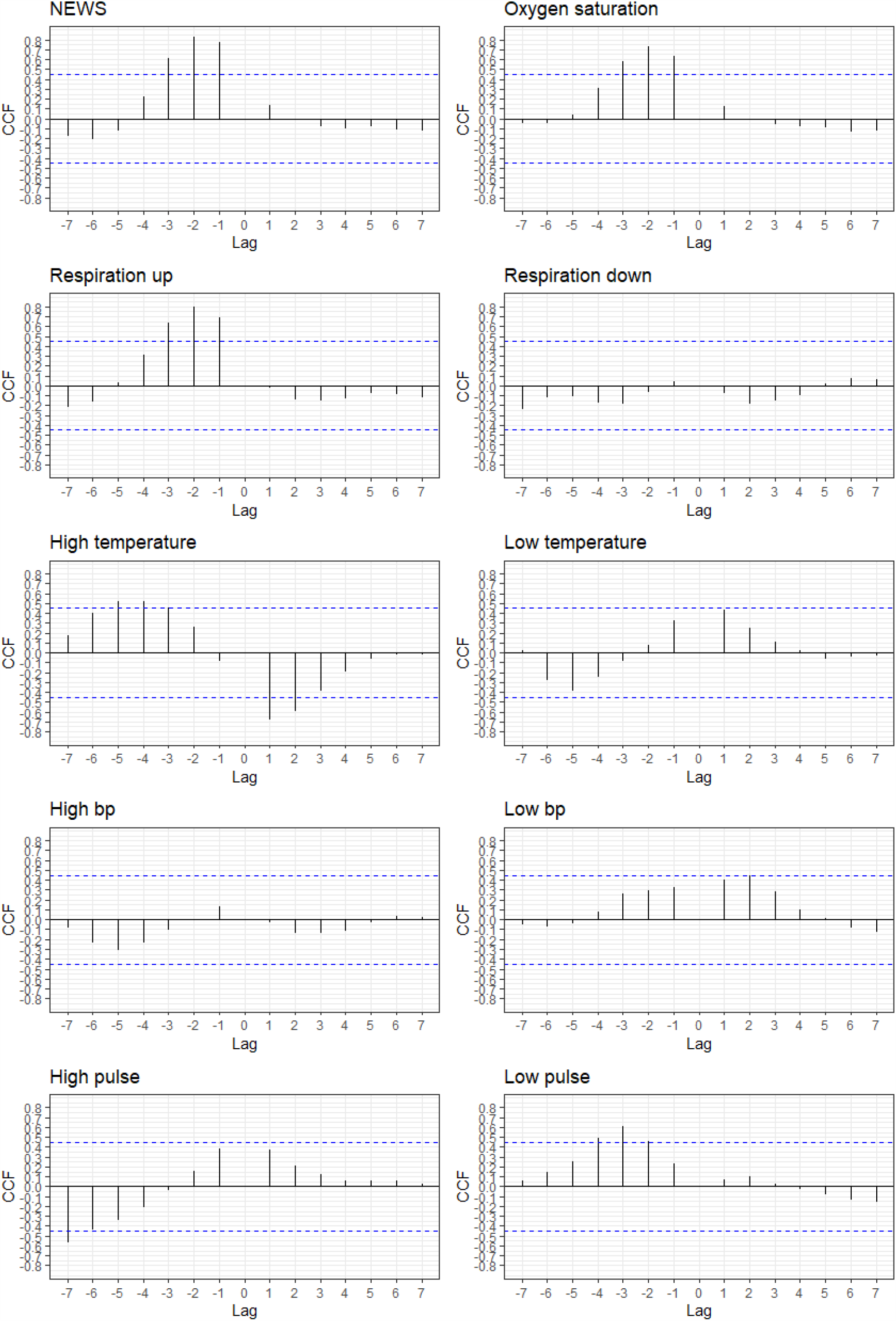
Cross-correlation function plots for NEWS (and component values) versus all cause deaths in care homes in matched geographical areas

## References

1. Comas-Herrera A, Fernandez J-L. England: Estimates of mortality of care home residents linked to the COVID-19 pandemic. International Long-Term Care Policy Network; 2020. [Available from: https://ltccovid.org/2020/05/12/estimates-of-mortality-of-care-home-residents-linked-to-the-covid-19-pandemic-in-england/ Accessed 2020-05-19]

2. Tay HS, Harwood R. Atypical presentation of COVID-19 in a frail older person. Age and Ageing. 2020.

3. Royal College of Physicians. National Early Warning Score (NEWS) 2 2020 [Available from: https://www.rcplondon.ac.uk/projects/outputs/national-early-warning-score-news-2 Accessed 2020-05-26]

4. British Geriatrics Society. COVID-19: Managing the COVID-19 pandemic in care homes for older people: good practice guide (last updated 14 April 2020) 2020 [Available from: https://www.bgs.org.uk/resources/covid-19-managing-the-covid-19-pandemic-in-care-homes#_edn5 Accessed 30/04/2020]

5. Scott LJ, Redmond NM, Tavaré A, et al. Association between National Early Warning Scores in primary care and clinical outcomes: an observational study in UK primary and secondary care. British Journal of General Practice. 2020:bjgp20×709337.

6. Barker RO, Stocker R, Russell S, et al. Distribution of the National Early Warning Score (NEWS) in care home residents. Age and Ageing. 2019;49(1):141–5.

7. Hodge S, Thompson C, Gordon AL. National early warning scores in care homes: do policy imperatives reflect a genuine need? Age and Ageing. 2019;49(1):5–6.

8. Smith GB, Prytherch DR, Schmidt PE, et al. Should age be included as a component of track and trigger systems used to identify sick adult patients? Resuscitation. 2008;78(2):109–15.

9. Greenhalgh T, Treadwell J, Burrow R, et al. NEWS (or NEWS2) score when assessing possible COVID-19 patients in primary care? Oxford: The Centre for Evidence-Based Medicine; 2020. [Available from: https://www.cebm.net/covid-19/should-we-use-the-news-or-news2-score-when-assessing-patients-with-possible-covid-19-in-primary-care/ Accessed 2020-05-21]

10. Whzan Digital Health. Early warning health detection systems 2020 [Available from: https://www.whzan.uk/ Accessed 2020-05-20]

11. Office of National Statistics. Deaths involving COVID-19 in the care sector, England and Wales: deaths occurring up to 1 May 2020 and registered up to 9 May 2020 (provisional): ONS; 2020 [Available from: https://www.ons.gov.uk/peoplepopulationandcommunity/birthsdeathsandmarriages/deaths/articles/deathsinvolvingcovid19inthecaresectorenglandandwales/deathsoccurringupto1may2020andregisteredupto9may2020provisional Accessed 2020-05-18]

12. R Core Team. R: A language and environment for statistical computing. Vienna, Austria: R Foundation for Statistical Computing; 2020.

13. Chen T, Wu D, Chen H, et al. Clinical characteristics of 113 deceased patients with coronavirus disease 2019: retrospective study. BMJ. 2020;368:m1091.

14. Verity R, Okell LC, Dorigatti I, et al. Estimates of the severity of coronavirus disease 2019: a model-based analysis. The Lancet Infectious Diseases.

15. Finnikin S, Hayward G, Wilson F, Lasserson D. Are referrals to hospital from out-of-hours primary care associated with National Early Warning Scores? Emergency Medicine Journal. 2020;37(5):279.

16. Scott LJ, Redmond NM, Garrett J, et al. Distributions of the National Early Warning Score (NEWS) across a healthcare system following a large-scale roll-out. Emergency Medicine Journal. 2019;36(5):287.

17. Brangan E, Banks J, Brant H, et al. Using the National Early Warning Score (NEWS) outside acute hospital settings: a qualitative study of staff experiences in the West of England. BMJ Open. 2018;8(10):e022528.

18. Silcock DJ, Corfield AR, Gowens PA, Rooney KD. Validation of the National Early Warning Score in the prehospital setting. Resuscitation. 2015;89:31–5.

19. Inada-Kim M, Knight T, Sullivan M, et al. The prognostic value of national early warning scores (NEWS) during transfer of care from community settings to hospital: a retrospective service evaluation. BJGP Open. 2020:bjgpopen20×101071.

20. Graham NSN, Junghans C, Downes R, et al. SARS-CoV-2 infection, clinical features and outcome of COVID-19 in United Kingdom nursing homes. medRxiv. 2020:2020.05.19.20105460.

